# Responsivity of the striatal dopamine system to methylphenidate – a within-subject I-123-ß-CIT-SPECT study in children and adolescents with Attention-Deficit/Hyperactivity Disorder

**DOI:** 10.1101/2021.11.19.21265993

**Authors:** Hans-Christoph Aster, Marcel Romanos, Susanne Walitza, Manfred Gerlach, Andreas Mühlberger, Natalie Hasenauer, Philipp E. Hartrampf, Kai Nerlich, Christoph Reiners, Reinhard Lorenz, Andreas K. Buck, Lorenz Deserno

## Abstract

**Background:** Methylphenidate (MPH) is the first-line pharmacological treatment of attention-deficit/hyperactivity disorder (ADHD). MPH binds to the dopamine (DA) transporter (DAT), which has high density in the striatum. Assessments of the striatal dopamine transporter by single positron emission computed tomography (SPECT) in childhood and adolescent patients are rare but can provide insight in how effects of MPH affect DAT availability. The aim of our within-subject study was to investigate the effect of MPH on DAT availability and how responsivity to MPH in DAT availability is linked to clinical symptoms and cognitive functioning.

**Methods:** Thirteen adolescent male patients (9-16 years) with diagnosis of ADHD according to DSM-IV and long-term stimulant medication (for at least 6 months) with MPH were assessed twice within 7 days using SPECT after application of I-123-ß-CIT to examine DAT binding potential (DAT BP). SPECT measures took place in on and off-MPH status balanced for order across participants. A virtual-reality continuous-performance test was performed at each time point. Further clinical symptoms were assessed for baseline off-MPH.

**Results:** On-MPH status was associated with a highly significant decrease (−27,6%) of striatal DAT BP as compared to off-MPH (t=4.93, p<0.001). More pronounced decrease in striatal DAT BP was associated with higher off-MPH attentional and externalizing symptom ratings (Pearson r=0.68, p=0.01). Striatal DAT BP off-MPH, but not on-MPH, was associated with higher symptom ratings off-MPH (Pearson r=0.56, p=0.04). In further exploratory analysis in left vs. right striatal sub-regions, stronger decrease in DAT BP in the right caudate nucleus was weakly associated with improved performance in the continuous-performance test (Pearson r= - 0.54, p=0.07).

**Conclusion:** Our findings corroborate previous reports from mainly adult samples that MPH reduces striatal DAT BP availability and suggest higher off-MPH DAT BP, likely reflecting low baseline DA levels, as a marker of symptom severity. More speculatively, regional specific responsivity of DAT BP to MPH may reflect treatment response with respect to cognitive functioning. However, implications from this small patient sample should be treated with caution and warrant replication.

## Introduction

Attention-deficit/hyperactivity disorder (ADHD) is a common child and adolescent psychiatric disorder associated with substantial psychosocial impairment on the individual and family level (1). ADHD significantly impacts on mental health and the social services (1). The clinical phenotype is characterized by motor hyperactivity, increased impulsivity and impaired attention starting in the most cases in childhood. There is a relevant increase of comorbid symptoms and disorders, for example social oppositional and conduct problems, substance use and depression (2). Clinically relevant ADHD symptoms persist into adulthood to a large extent (3).

Pharmacological interventions by stimulant medication, such as methylphenidate (MPH), are effective and save (4). MPH binds primarily to the dopamine and to some extent to noradrenaline transporter and because of the high density of the dopamine transporter (DAT) in the striatum, striatal DAT is one of MPH’s main neurochemical targets (5) and may thus be related to MPH treatment response. Considering different response criteria the response to MPH is limited to approx. 70% of childhood patients (6), the identification of the neurobiological correlates (or even determinants) may allow to establish quantitative markers of treatment response in the long run to provide more individualized outcome predictions in the future. Therefore, the investigation of the striatal dopamine system by means of Single-Positron-Emission-Computed Tomography (SPECT) and Positron Emission Tomography (PET), and in particular its modulation by stimulant medication in an children’s and adolescents population, may provide a better understanding of whether MPH-related changes in DAT availability are indeed related to changes in clinical symptoms.

Numerous functional magnetic resonance imaging (fMRI) and Electroencephalography (EEG) studies have been published on altered neurocognitive processes in fronto-striatal brain circuits of ADHD patients for example during executive function (7, 8), Studies approaching the striatal dopaminergic system by SPECT or PET are clearly more challenging to conduct, especially in an children’s and adolescents population. However, molecular imaging techniques can provide insights to neurochemical targets of a drug (9). Due to the ethical limitations and high costs of neurochemical imaging studies with radioactive ligands, only few studies with relatively small numbers of patients were published until now (10). These studies are highly heterogeneous in regard to sample characteristics as well as the applied tracers rendering direct comparisons considerably difficult, especially when results are conflicting. Nonetheless, DAT has been studied most consistently as summarized in a meta-analysis of 9 PET and SPECT studies in adult patients (10). Adult patients with ADHD had a higher striatal DAT availability (likely indicating lower striatal DA levels (11)). However, the study showed that about half of the variability in DAT availability was related to medication status. Patients who were already treated with psychostimulants, such as MPH, showed a significantly higher DAT availability than groups of patients who were drug-naïve (10). This was discussed as an adaptive response to the long-term blockade of dopamine transporters by continuous treatment. This could in turn explain the partial decrease in long-term efficacy of MPH at the same dose. However, only cross-sectional studies were included (10), which limits conclusions in this regard. A longitudinal study, however, confirmed that one year treatment on MPH in adult ADHD lead to enhanced DAT availability (12). Nonetheless, when using a tracer that binds to D2 receptor to measures displacement by one-year MPH treatment as marker of DA levels in adult ADHD, it was found that MPH treatment lead to elevated DA levels (13). This increase in DA levels (measured via D2 receptor displacement) was correlated with improved inattention symptoms after one-year MPH (13).

Although the dopaminergic system undergoes substantial development changes (14, 15), to our knowledge only two studies investigating the effect of MPH therapy on DAT have been published in adolescent ADHD. Results from both studies indicate that MPH treatment leads to a decrease in DAT availability in response to MPH treatment (16). However, only one of the two studies found a relation to clinical symptoms based on a subgroup analysis. Neither study reported on relationships with neurocognitive performance measures. To fill this gap, we here detail a 123-ß-CIT SPECT study on 13 male children and adolescents with ADHD, who were on continuous medication (for at least 6 months) on MPH before study enrollment. All patients performed two SPECT measurements (counter-balanced for order), one on MPH, and another one after 7 days of treatment cessation (off-MPH). Binding of 123-ß-CIT is aiming at quantifying DAT density 24h after injection. We concomitantly applied both clinical rating scales as well as neurocognitive testing procedures in a virtual reality setting in order to determine whether DAT density was correlated to measures of ADHD symptoms and cognitive performance.

## Methods

### Study design

All participants were on continuous MPH medication for at least 6 months prior to inclusion in the study. During this period, MPH was taken once a day between 7 and 8 a.m. by oral administration. Patients received daily doses of MPH between 0.4 and 1.2 mg/kg body weight. Those dosage regimes were met individually based on clinical needs and prescribed by the respective treating child and adolescent psychiatrists. All participants received long-acting MPH preparations (Medikinet retard® or Concerta®) ensuring continuous efficacy of the treatment during the lengthy procedures of the study. The cessation phases required for participation were organized to comply with clinical demands such as regular drug holidays or yearly cessation to determine further necessity of the drug. In all patients, treatment was reintroduced after study participations based on the families and the physicians estimate that medication provided significant benefit. The study procedure was accomplished on four days. Day A and B were two consecutive days as were day C and D with one week in between. Investigations on day A and B were performed on MPH, on day C and D off-MPH. To counter-balance for order effects, 9 patients were investigated first on MPH, and then again after 7 days of cessation while 4 patients initially stopped medication for 7 days before the first SPECT measurement and were assessed a second time after 7 days of MPH treatment. Study compliance was assessed by self- and mother’s report.

### Sample

The study was approved by the Federal Office for Radiation Protection (“Bundesamt für Strahlenschutz”) and the ethics committee of the University of Wuerzburg and was performed in accordance with the ethical standards laid down in the latest version of the Declaration of Helsinki. 13 male children and adolescent patients with diagnosis of ADHD according to DSM-IV and aged 9-16 years (mean ± S.D.: 11.3 ± 2.8) were recruited in the outpatient clinic of the Department of Child and Adolescent Psychiatry, Psychosomatics and Psychotherapy of the University of Wuerzburg. All patients and their legal guardians gave written informed consent before inclusion in the study. All participants were thoroughly assessed by complete semi-structured interview (Kiddie-Sads-PL, German version) determining ADHD status and comorbid disorders. Parents filled in the Child behavior Checklist and the “Depressionsinventar für Kinder-und Jugendliche” determining depressive symptoms. Exclusion criteria were female, sex, history of birth complication, traumatic head injury, seizures, severe somatic or neurological disorders, psychosis, autism spectrum disorder, severe conduct disorder, and IQ below 85. All patients had received continuous pharmacological treatment with MPH for at least 6 months before inclusion in the study (see Table 1 for sample characteristics).

### Single Photon Emission Computed Tomography

To assess the DAT availability and binding potential, respectively, the highly affine presynaptic dopamine transporter ligand, Iodine-123-β-CIT [2-β-carboxy-methoxy-3-β-(4-iodophenyl)-tropane] was used (17, 18). Image acquisition was performed on Day B and D 24 h after i.v. injection of weight-adapted activity of Iodine-123-β-CIT on Day A and C (19) with a dual-head gamma camera (E.cam duet; Siemens) fitted with middle-energy, parallel-hole collimators. Energy window for iodine-123 was set on 160 keV +/- 15%. For each scan, 60 projections (40s per frame) were collected in step-and-shoot mode on a circular orbit. The image data were reconstructed by standard filtered back projection using Butterworth filter with cut-off frequency of 0,6/cm and order 8, followed by attenuation correction using a first-order method (20). To block binding of free iodine in the thyroid gland, the patients took 16,4mg/kgKG sodium perchlorate (Irenat) orally 20 minutes before injection of I-123-β-CIT, followed by 6mg/kgKG Irenat three times a day the following three days. For evaluation (intra-individual comparison of the influence of methylphenidate treatment) and standardized quantification by stereotactic normalization patient studies were co-registered with a normal template of a healthy age matched control subject. Standardized templates (in size and shape) created in our normal template (based on Colin 27 NMR-Brain (21)) were used for defining the striatal regions of interest. After transforming the region of interests (ROI) templates for the striatum and its subregions as well as for the reference region in the occipital lobe from the normal template onto the two SPECT/CT patient studies of interest (on/off MPH), the position of the ROI templates were manually adjusted to fit individuals because of anatomical differences (Figure 1). We evaluated the data derived from three consecutive transverse slices, which showed the highest tracer accumulation in the basal ganglia. For semi-quantitative evaluation, DAT availability was calculated using the binding potential (22) ratios [(STR-BKG)/BKG] in the striatum (STR) and its subregions (putamen, caudate nucleus) with an occipital reference region used as background (BKG), as recommended by EANM Guidelines (23) and already implemented in this way in other previous studies (24).

**Figure 1:**
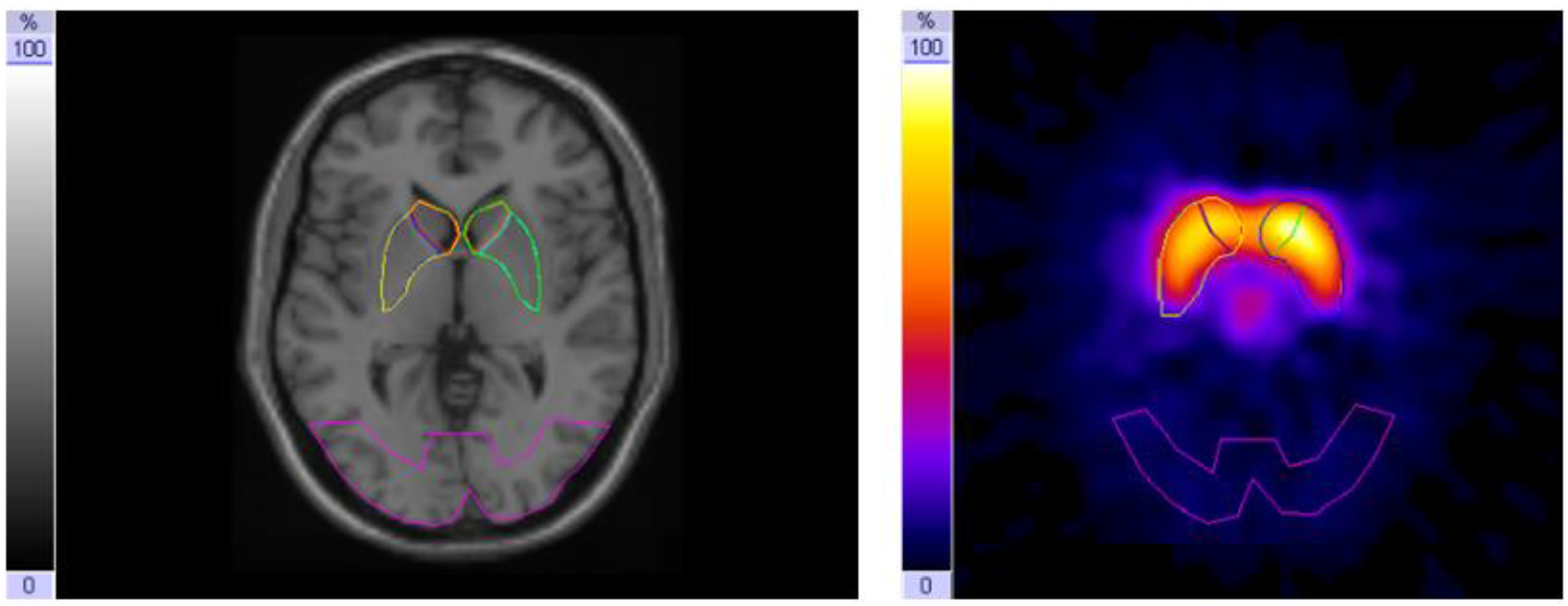
MRI with ROI-template-set covering the striatum with caudate nucleus and putamen and an occipital background ROI (left); I-123-ß-CIT-SPECT (summation of the three consecutive slices with the highest tracer uptake in the basal ganglia (right))

### Continuous performance test in virtual reality classroom

On Day A and Day C, patients performed a neurocognitive testing procedure in a Virtual Reality Classroom (VRC) (25, 26) thus assessing the participants’ attention in a more ecological valid setting. Within the VRC, a continuous performance test (CPT) was presented on a blackboard while distracting visual (e.g., another teacher coming into the room) and auditory (e.g., coughing classmate) stimuli appear. Participants’ head movements were measured. The VR environment and instructions were presented with a Z800 3D Visor head-mounted display (HMD; eMagin, Hopewell Junction, USA) and a closed headphone (Sennheiser, Wedemark, Germany), both connected to a notebook with Microsoft Windows operating system. The head position was monitored by an electromagnetic tracking device (Minuteman, Polhemus Corporation, Colchester, USA) in order to adapt the field of view to head movements and to assess head orientation.

The experimenter explained the VRC equipment, made sure the child’s eyesight, hearing and cognitive capabilities were unimpaired and adjusted the HMD to the child’s head, so that the interior of the virtual classroom could be seen. The child then was instructed to look around in the room to adapt to the virtual environment. The child was then given the first task (warm-up task) by the virtual teacher. It was instructed to view a series of numbers appearing on the blackboard and to hit the button of the responding device every time the number “5” preceded by the number “9” occurred. Each of the numbers remained on the blackboard for 200 milliseconds (ms) followed by an interstimulus interval of 800 ms. Altogether twenty number stimuli were presented containing five target sequences. If necessary instructions and warm-up trial were repeated. In case the task was completed without making more than one error, the second task (main task) started after a short break.

In the main task the participant was instructed by a virtual teacher to view a series of letters presented on the blackboard. The participant had to press the response button as quickly as possible every time he detected the letter “K” preceded by the letter “A”. Altogether two blocks of each 680 letter stimuli including 72 target stimuli were presented with a stimulus duration of 200 ms and an interstimulus interval of 800 ms. The first block included no distracters. Directly after the first block a second block including the same stimuli and target number was presented. This block included pure auditory (typical classroom sounds like whispering, dropping pencils, etc.), pure visual (paper plane flying across the room) or mixed auditory/visual (bus noisefully driving by outside left window) distracters distributed over the whole block. Each block lasted 11 minutes and 20 seconds.

### Child Behavior Checklist (CBCL)

The CBCL in the version for children and adolescents aged 4-18 years captures parents’ assessments of their children’s competencies and problems. The evaluation of this questionnaire includes the following scales and scores: 3 competency scales (activity, social skills, and school), 8 cross-assessment syndromes (Social Withdrawal, Physical Discomfort, Anxiety/Depressive, Social Problems, Schizoid/Compulsive, Attention Deficit Disorder, Dissocial Behavior, Aggressive Behavior) where comparison is possible across parent, teacher, and self-report forms of this questionnaire system. Scales of internalizing and externalizing behavior and an overall problem behavior score are formed from the syndrome scales (27, 28). The questionnaire was completed by the patients’ parents at the time off-MPH.

### Statistical Analysis

The statistical analyses were conducted using the JASP Toolbox (JASP Team (2020, Version 0.14.1). Specific DAT binding of the patients on and off-MPH 24h after administration of 123-ß-CIT was compared using a generalized linear mixed effects model with medication status as fixed effect and individual subjects and medication order as random effects (striatal binding potential ∼ medication status + (1|participant) + (1|order). Change scores in regional binding potential and performance in the CPT were calculated by the difference of the respective values at the time of off medication minus on medication. The influence of MPH on the performance in the virtual classroom setting was also analyzed using a generalized linear mixed effects model (correct choices ∼ medication status + (1|participant) + (1|order). For correlations, after testing for a normal distribution using the Shapiro-Wilk test, we used Pearson correlation coefficients. All variables were z-scored before analysis. Comparison of correlation coefficients, when indicated, were calculated by a Fisher’s z-transformation (29).

## Results

For a sample description, see Table 1.

### Single Photon Emission Computed Tomography

In a linear mixed effects model with the individual participants as random effects, there was a significant difference in striatal binding potential (BP) between the on and off-MPH DAT BP (degrees of freedom: 20.6, t-value: 4.93, p-value < 0.001, beta estimate: -2.68 (SD: 0.54), see Figure 2 and 3). The medium reduction of DAT binding potential was 27,6 % (median 32.1%), ranging from -7.4% to 68.9% (SD: 19.4%). Nominal reductions in the medication condition vs. the non-medication condition were found in all participants except for one, who showed basically no change in DAT and a second participant, who displayed a nominal increase of DAT.

**Figure 2:**
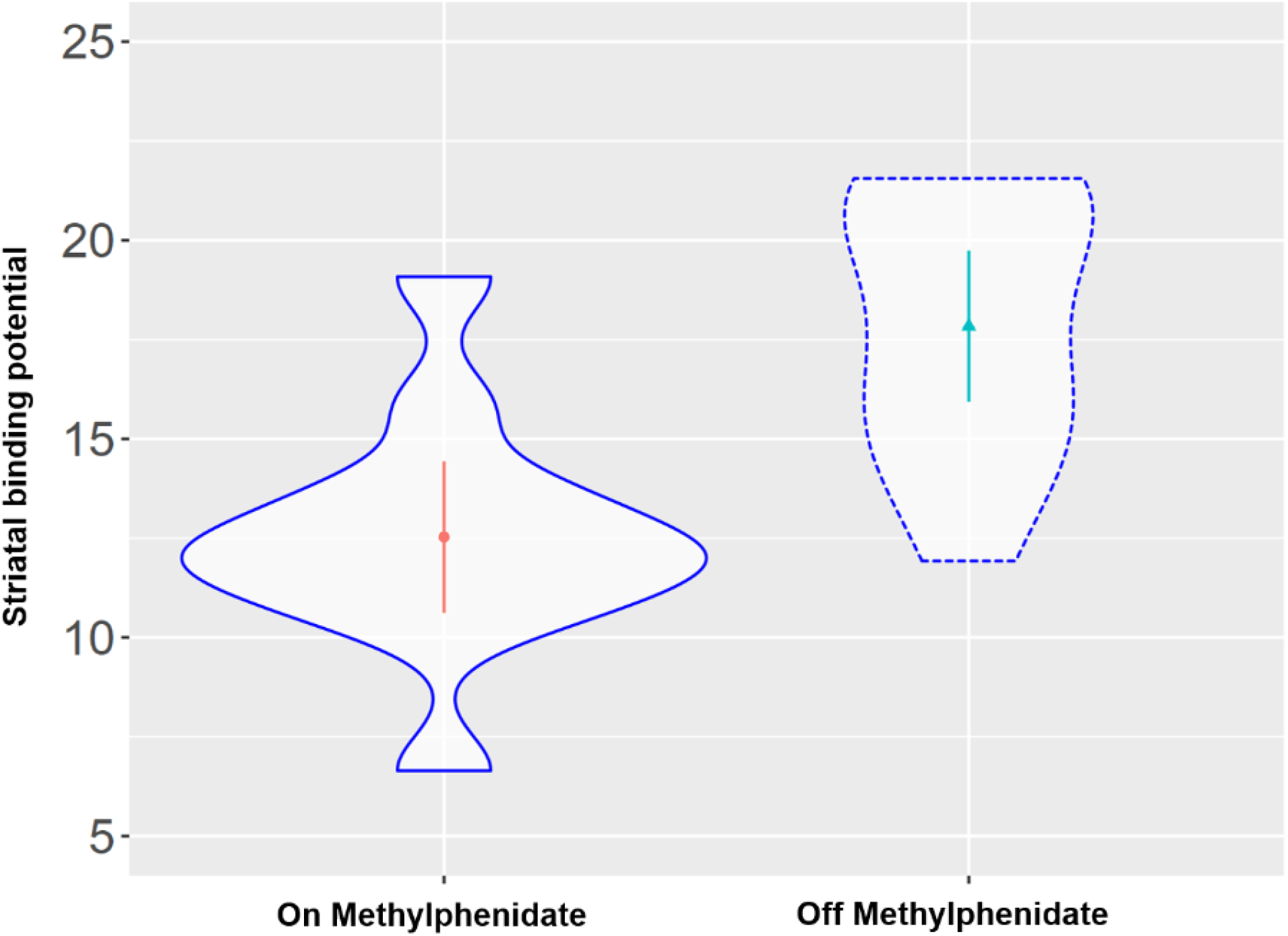
Comparison of striatal DAT binding potential on and off-methylphenidate (p < 0.001).

**Figure 3:**
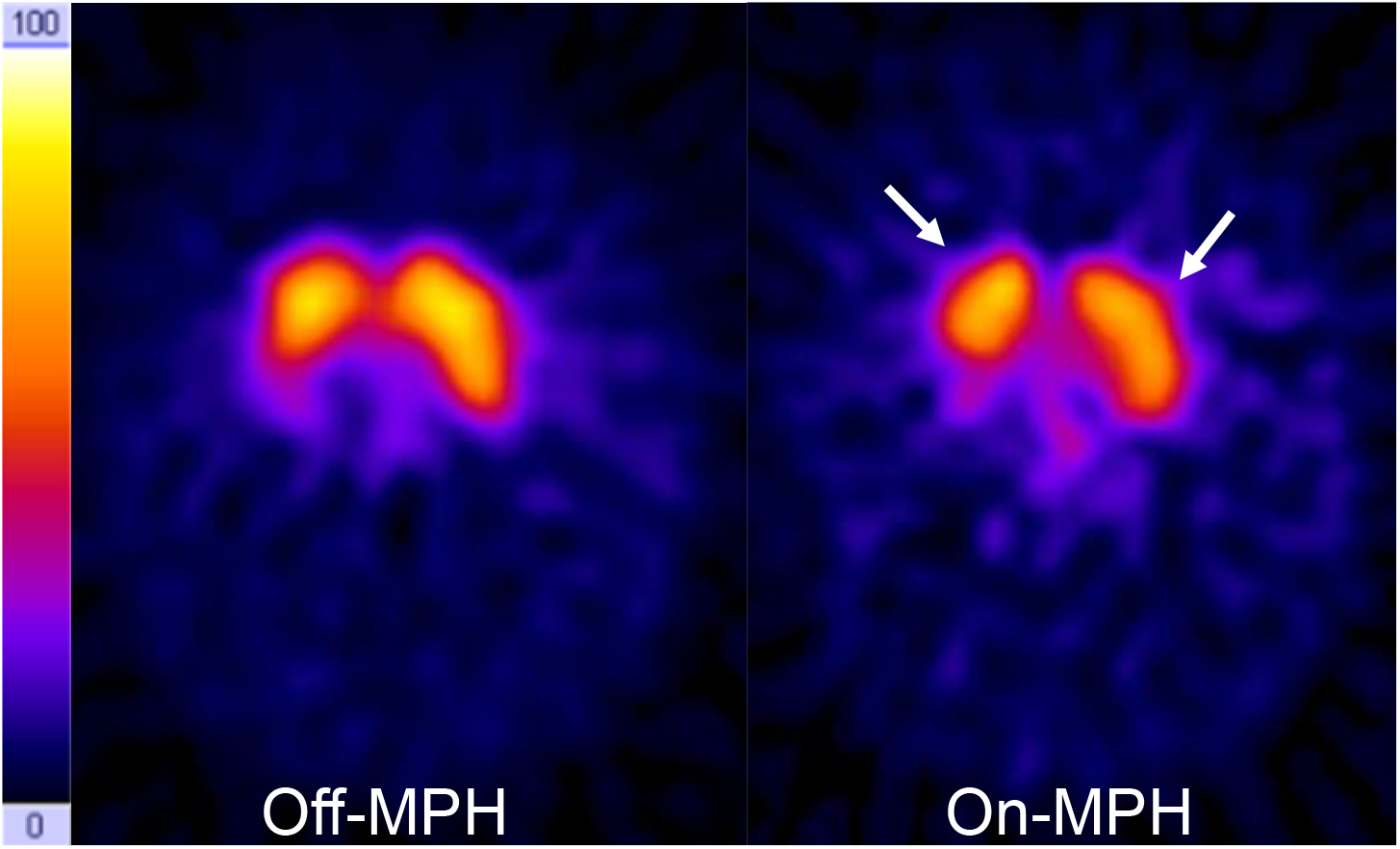
DAT-scans on and off-MPH in a single subject. Acquisition time points were 24h after tracer application. Color sale is hot metal. Time span between the two images was seven days. On the right side a decrease of tracer accumulation in the basal ganglia can be seen (arrows).

### Continuous Performance Task

In a mixed linear effects model with medication status as a fixed effect and participants as a random effect showed no significant effect on the continuous performance task parameters (correct choice (Figure 4), errors and head movement (degrees of freedom: 11, t-value: 0.74, p-value: 0.47, estimate: 0.04 (SD:0.06))).

**Figure 4:**
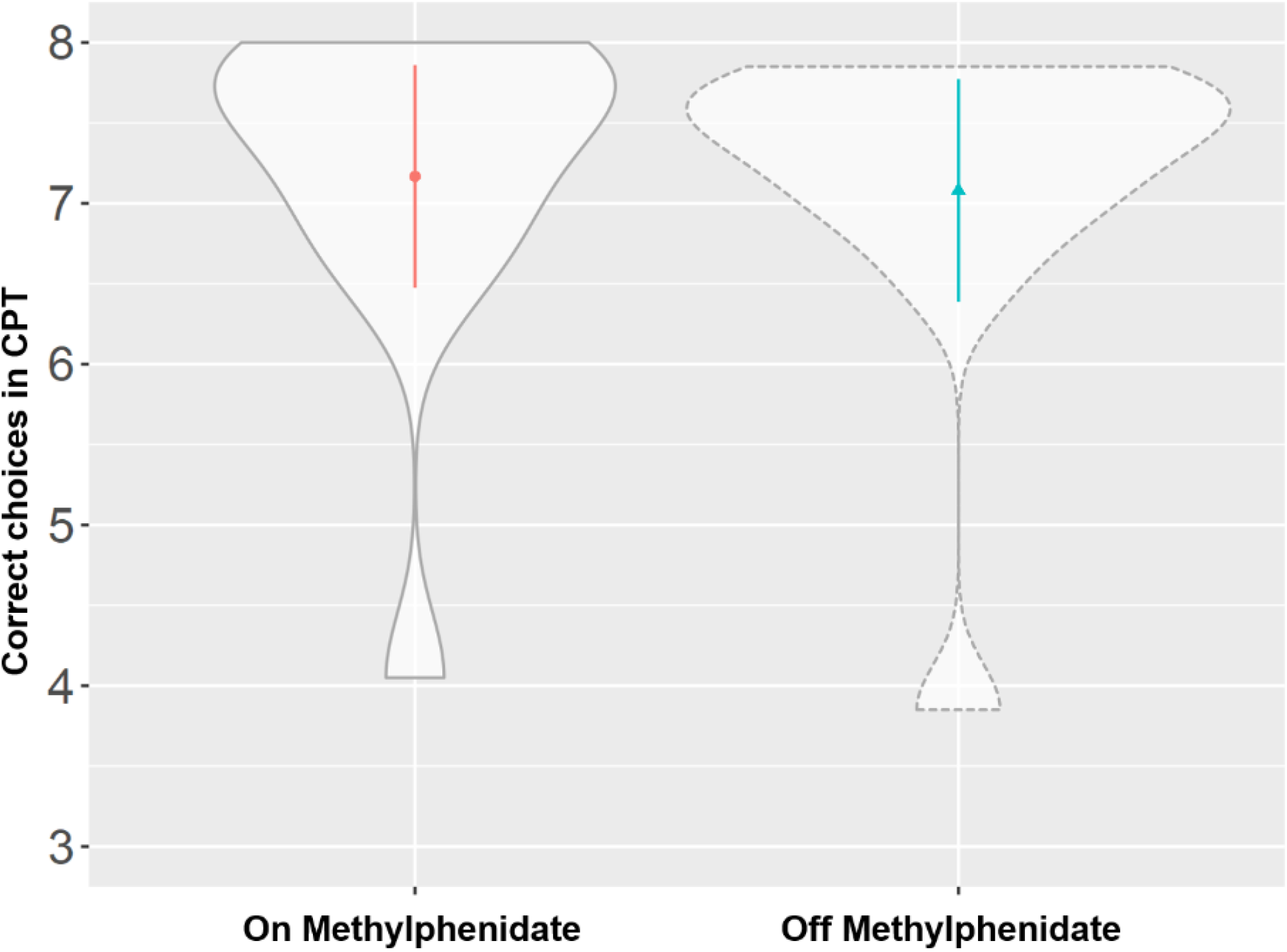
Comparison of correct choices in the continuous performance task on and off-MPH (p-value: 0.47).

### Correlations between change of binding potential and clinical measures

The parent-rated severity of internalizing and externalizing symptoms was evaluated at the time off-MPH using the CBCL questionnaire. The severity of externalizing symptoms correlated with the change in binding potential in the striatum due to MPH medication (Pearson r=0.68, p=0.01, Figure 5). This effect was driven by the level of striatal binding potential off MPH (Pearson r=0.56, p=0.04) and not by the striatal binding potential on MPH (Pearson r=-0.36, p=0.22). The severity of internalizing symptoms was neither correlated with the change in striatal binding potential (Pearson r=0.47, p=0.11) nor the striatal binding potential off (r=0.43, p=0.19) and on (r=-0.19, p=0.54) MPH.

**Figure 5.**
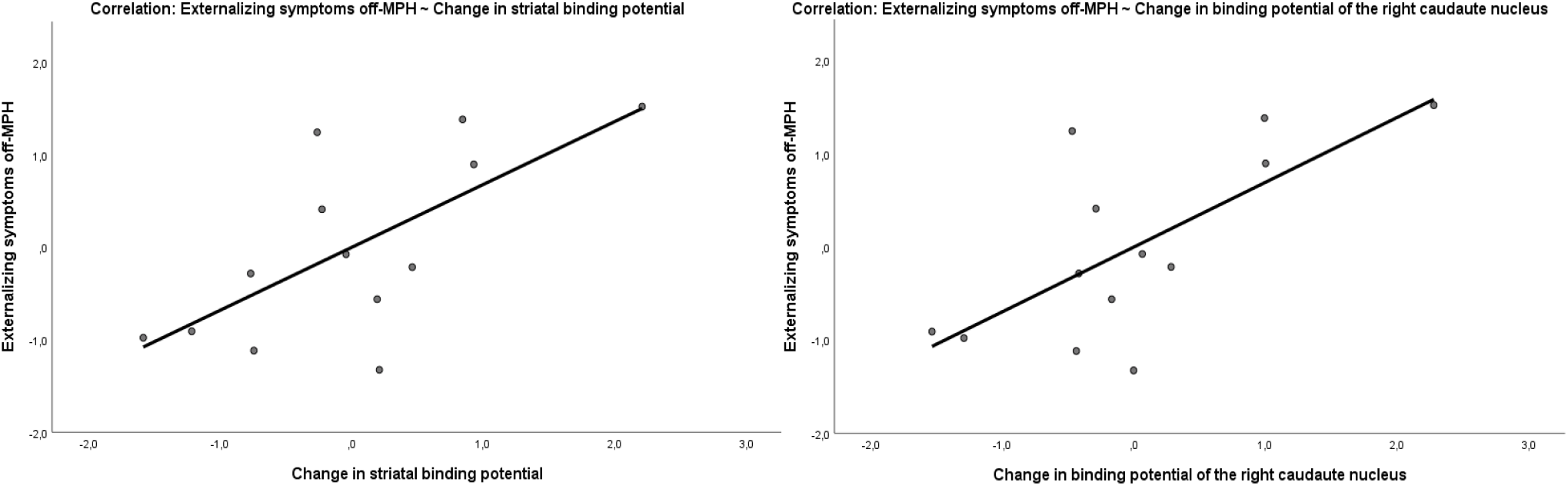
Correlation of the externalizing symptoms score off MPH with the change of binding potential in the striatum (Pearson r=0.68, p=0.01) and in the right caudate nucleus (Pearson r=0.7, p=0.008).

To explore potential regional and hemisphere specific effects in the striatum linked to symptom severity, the binding potentials of the left and right caudate nucleus and the putamen were estimated separately. In a first step, we compared the correlation coefficients of the change in the binding potential of the caudate nucleus (Pearson r=0.74, p=0.004) and the putamen (Pearson r=0.47, p=0.1) with externalizing symptom severity, which differed significantly (z=-1.99, p=0.04). Between the hemispheres, no relevant differences in correlation strength were found. The change of the binding potential in the left putamen (r=0.63, p=0.02) had a stronger, but not significantly stronger (z=-1.79, p=0.09), correlation with externalizing symptoms than the change of the binding potential of the right putamen (r=0.17, p=0.58). The change of the binding potential of right caudate nucleus (Pearson r=0.7) showed only a slightly stronger correlation with the externalizing symptoms score than the correlation of the left caudate nucleus (Pearson r=0.65) with the externalizing symptoms score, which was not significant (z=0.5182, p-value=0.6043). The strongest correlation with symptom scores and change in binding potential was seen in the right caudate nucleus. Change in binding potential of the right caudate nucleus correlated with the externalizing (Pearson r=0.7, p=0.008) (Figure 5), but also with the internalizing (r=0.58, p=0.04) symptom severity of the patients at the time off-MPH.

### Correlations of changes in CPT measures and binding potentials

Because the CPT was performed twice within subject on and off MPH, change scores could also be calculated in this case. The change in striatal binding potential did not correlate with a change in CPT outcomes (correct choices (Pearson r=-0.45, p=0.14), errors (Pearson r=0.34, p=0.28), reaction times (Pearson r=0.09, p=0.78), head movement (Pearson r=-0.42, p=0.19)).

In explorative analyses, there was a marginally non-significant correlation of the change in the binding potential of the right caudate nucleus with the change in the correct responses in the CPT (Pearson r= - 0.54, p=0.07). Because the change scores of the binding potentials and the CPT scores were calculated off-on MPH, a better performance in the CPT under medication implies a decrease in the change score, therefore the correlation of the change of the binding potential and the correct responses in the CPT is negative (Figure 6). To test whether this effect was specific to the right caudate nucleus, the correlation coefficients of the change in correct responses in the CPT and binding potentials in the right putamen and left caudate nucleus were compared with those of the right caudate nucleus. There was a significant difference in the strength of the correlation between the right caudate nucleus and the right putamen (z = -2.5917, p-value = 0.009) and a marginal difference in the correlation strength between the left and right caudate nucleus (z = 1.9124, p-value = 0.05). The change in binding potential in the right caudate nucleus did not correlate with any other outcome change in the CPT (errors (Pearson r= 0.38, p=0.23), head movement (Pearson r=-0.31, p=0.36) and reaction time (Pearson r=0.1, p=0.75)). The change of binding potential of the right (r=0.22, p=0.49) and left (r= -0.47, p=0.12) putamen, or the left caudate nucleus (r= -0.32, p=0.32) did not correlate with the change of correct responses in the CPT.

**Figure 6.**
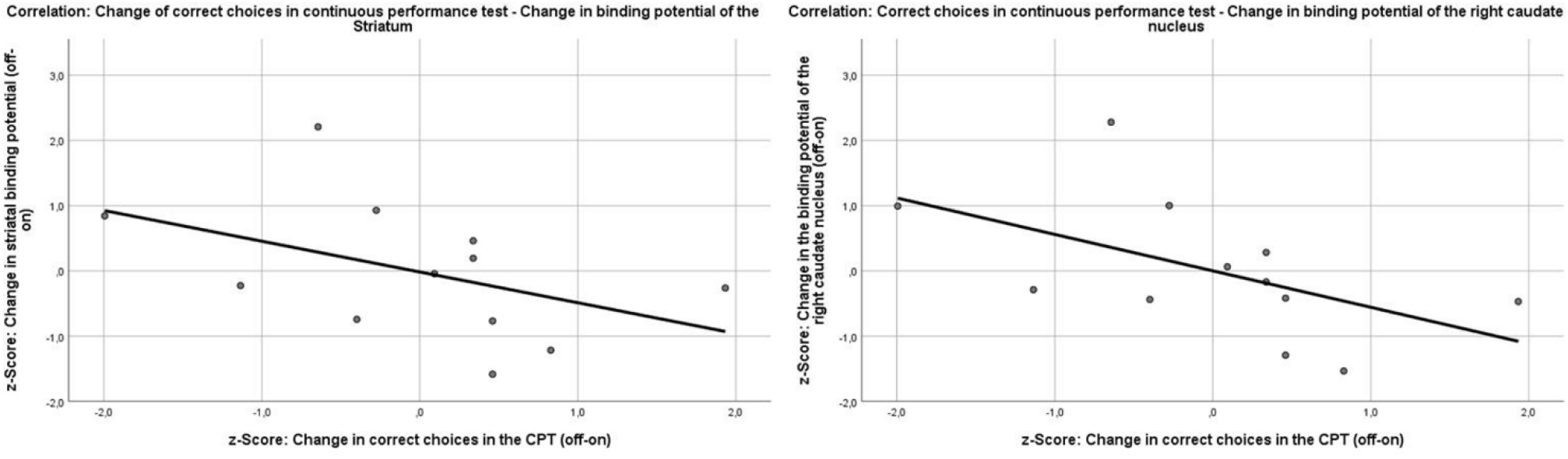
Correlations between the change of the binding potentials in the striatum (Pearson r=-0.45, p=0.14) and the right caudate nucleus (Pearson r= - 0.54, p=0.07) and the change of correct choices in the continuous performance test.

## Discussion

We present one of few within-subject studies assessing striatal DAT density in children and adolescents with ADHD. Our results show that MPH administration leads to a significant decrease in DAT availability. The severity of externalizing symptoms off-MPH correlated with the change in DAT. Further explorative analyses indicated a correlation between change in DAT in the right caudate nucleus and change in the correct choices in the CPT. Especially correlations with symptoms and CPT thus add substantially to the small body of literature. Further, our study extends previous studies by the type of design.

Our data corroborate the observation from previous studies that MPH on a group level leads to a significant reduction of DAT binding (likely reflecting higher striatal DA levels). Despite this already established finding in adults and adolescents (5, 13, 30), our study is the first to actually provide evidence for a positive correlation between striatal DAT binding off-MPH and the severity of externalizing symptoms off-MPH. Region-specific analyses showed a marginally non-significant correlation between the decrease in DAT availability in the right caudate nucleus and the improvement in performance on the CPT. To our knowledge, previous studies in children or adolescents have not investigated this question (31, 32) or did not find significant correlations (24, 33). The study by Szobot et al. recruited psychostimulant-naive patients with substance abuse disorder including cannabis, cocaine and alcohol. Medication of MPH (1.2mg per kg body weight) for 3 weeks decreased core ADHD symptoms and striatal DAT availability. However, neither naive DAT availability related to clinical effect, nor were there correlations between changes in ADHD symptomatology and DAT availability (24). The study by Akay et al. included under-age patients over a 2-month period to evaluate the effects of MPH on striatal DAT availability and ADHD core symptomatology. Therapy with MPH lead to reduced striatal DAT availability (and ADHD core symptomatology). In the latter study, a subgroup comparison showed that the patients with significantly reduced ADHD core symptomatology by MPH, also showed a greater decrease in DAT availability (16). However, all patients included in our studies received long-term MPH treatment, which has been discussed to lead to an upregulation of DAT in adaption to long-term MPH treatment (10).

Both of the previous studies used 99mTc-TRODAT-1 as radioligand to quantify active dopamine transporters in the striatum, whereas our study used I-123-ß-CIT as radioligand. In a study with Parkinson’s disease patients, I-123-ß-CIT was found to be superior in sensitivity and specificity for quantifying the functionality of dopaminergic synapses (34). Furthermore, both studies recruited only psychostimulant-naive patients, whereas the patients in our study were all long-term treated with MPH before study inclusion. This may have had an effect on the availability of DAT off-MPH (10, 12, 13). Our study results are in line with meta-analytic evidence from genome-wide association studies for an association between the genes encoding DAT (SLC6A3), Catechol-O-Methyltransferase (COMT) or the dopamine receptor D4 (DRD4) and the clinical efficacy of MPH (35). MPH and amphetamines, which both act mainly at DAT, are established as the most effective medication as first-line therapy (4). A large number of cognitive neuroimaging studies have shown alterations in the dopaminergic mesolimbic system, a neural circuit associated with motivated behaviors, reward learning and higher-order cognition (36-38). One hypothesis is that extracellular dopamine levels and tonic release of dopamine are decreased in ADHD patients, resulting in increased stimulus-dependent phasic releases of dopamine (potentially via presynaptic auto-receptors (39)). This impaired signal-to-noise ratio could provide increased distractibility to external stimuli (40). Another hypothesis is that both phasic and tonic release of dopamine is decreased in ADHD patients, which would explain why effects of reinforcement are weakened in ADHD patients to motivate behavior (41-43). Indeed, increased DAT availability could indicate lowered extracellular dopamine levels. In sum, our study is in line with relatively lower DA levels (higher DAT) before treatment but cannot contribute to the question how phasic DA release might be altered. However, also the factors contributing to decreased extracellular dopamine levels are still matter of ongoing research. For example, an overactivity of the DAT itself, a dysfunction of dopamine receptors or presynaptic autoreceptors are conceivable to play a role (44, 45). These effects could also emerge as a result of aberrant glutamatergic and GABAergic influences over the dopamine system (46).

The right caudate nucleus and its possible role in the pathophysiology of ADHD is already described in other publications. A study attempted to measure tonic release and phasic dopamine release with a D2 PET displacement measurement during a response inhibition task. They found that dopamine levels of ADHD patients, specifically in the right caudate nucleus, were generally lower but higher during the response inhibition task (putatively reflecting higher phasic release) as compared to healthy adults (47). While these changes were also visible throughout the striatum, they were significant in the right caudate nucleus (47). A larger PET study in adult ADHD patients also confirmed increased DAT availability in the right caudate nucleus of ADHD patients, which could be indicative of lower tonic dopamine levels (48). A meta-analysis comparing structural MRI images between ADHD patients and healthy controls showed that specifically the right caudate nucleus is one of the regions that differ most in volume from controls in ADHD patients (49). A meta-analysis of 23 fMRI found a significant difference between minor and adult ADHD patients exclusively in the right caudate nucleus. In this meta-analysis, activation in the right caudate nucleus was lower in minors than in adult ADHD patients during response inhibition tasks (50). A randomized clinical trial using a Go/NoGo task during fMRI to assess potential biomarkers for predicting clinical response to MPH showed increased baseline activation in the right caudate nucleus in patients with a particularly good clinical response to MPH, but only when considered in combination with response to atomoxetine (51).

One limitation of our study is that we provide intra-individual comparisons of pre-medicated patients in absence of a healthy control group. Thus, we cannot determine the effects of chronic medication on the status prior to installation of MPH in our participants. However, a meta-analysis on SPECT and PET studies investigating DAT binding in ADHD compared to controls suggests that a history of medication may lead to adaptation resulting in higher DAT binding (10). One may argue that in line with the meta-analysis by Fusar-Poli et al. DAT binding in our sample has already been increased due to chronic medication and that cessation of chronic MPH treatment may have further increased DAT availability. This increase in DAT availability over 12 months of MPH therapy has already been demonstrated in a prospective study (12). In line with this notion, secondary adaptation processes may reverse or at least weaken the initial pharmacological effect of reducing DAT availability. This has been suggested as an account of decreased efficacy of MPH treatment in the long run. Another limitation of our study design was that no on-drug clinical evaluations were collected to examine the clinical effect of MPH and correlate it with the changes in binding potentials. Only the continuous performance test was performed on and off drug. Given our small sample size of 13 patients, our results should be considered an exploratory analysis that requires replication with a larger sample also because sample sizes are prone to substantially overestimate effect sizes (52). This needs to be emphasized in particular because exploratory correlation analyses reported were not corrected for multiple comparisons.

In summary, the results of our study support the importance of the DAT, specifically in the striatum, in relation to responsivity to MPH. Our findings could inform future studies exploring individualized prediction of the treatment efficacy of MPH in childhood and adolescent patients. This potential has already been indicated by two other studies in adolescent (53) and adult (54) ADHD patients. Besides the established nuclear imaging methods, it would also be beneficial to test proxy markers of DA, and also monoamine function more generally, for example by making use of recently developed quantitative MRI methods (55). In combination with specifically designed neurocognitive tests targeting the interaction of cognition and motivation from a reinforcement learning perspective (56) this could provide an in-depth understanding of inter-individual differences in the treatment response to MPH to enable more individualized treatments in the future.

## Data Availability

All data produced in the present study are available upon reasonable request to the authors.

## Data Availability

All data produced in the present study are available upon reasonable request to the authors.

## Funding

This study was supported by a grant from the Interdisciplinary Centre of Clinical Research at the Medical Faculty of the University of Würzburg (“Untersuchungen des Dopamin-Transporters mittels TC-99m-TRODAT-1-SPECT bei Jugendlichen mit ADHS”) and by the German Research Foundation (DFG) as part of KFO 125 “Attention Deficit/Hyperactivity Syndrome (ADHD): Molecular Pathogenesis and Endophenotypes in the Course of Treatment”, project number 5397423. HCA is supported by a Clinician Scientist Program at the Interdisciplinary Centre of Clinical Research at the Medical Faculty of the University of Würzburg. LD is supported by the IFB Adiposity Diseases, Federal Ministry of Education and Research (BMBF), Germany, GN: 01EO150 and the German Research Foundation (DFG) as part of the Collaborative Research Centre 265 “Losing and Regaining Control over drug intake” (402170461, Project A02).

## Author contribution

HCA statistically analyzed the data and drafted the manuscript under supervision of LD. MR, SW, MG developed the study design, recruited patients and collected clinical data. AM collected and analyzed the CPT data. RL, CR, AB, PH, NH, KN were involved in SPECT data collection and analyzed SPECT binding potentials data. All authors read and revised the manuscript.

## Notes

### Competing Interest Statement

The authors have declared no competing interest.

### Author Declarations

The ethics commitee of the medical faculty of the University of Wuerzburg and the German Federal Office for Radiation Protection gave ethical approval for this work.

